# Sample Size Calculation for Phylogenetic Case Linkage

**DOI:** 10.1101/2020.07.10.20150920

**Authors:** Shirlee Wohl, John R. Giles, Justin Lessler

## Abstract

Sample size calculations are an essential component of the design and evaluation of scientific studies. However, there is a lack of clear guidance for determining the sample size needed for phylogenetic studies, which are becoming an essential part of studying pathogen transmission. We introduce a statistical framework for determining the number of true infector-infectee transmission pairs identified by a phylogenetic study, given the size and population coverage of that study. We then show how characteristics of the criteria used to determine linkage and aspects of the study design can influence our ability to correctly identify transmission links, in sometimes counterintuitive ways. We test the overall approach using outbreak simulations and provide guidance for calculating the sensitivity and specificity of the linkage criteria, the key inputs to our approach. The framework is freely available as the R package *phylosamp*, and is broadly applicable to designing and evaluating a wide array of pathogen phylogenetic studies.

## Introduction

As the cost of pathogen sequencing has declined, the number and size of studies based on pathogen sequence analysis has increased dramatically (Neher and Bedford 2018). Traditionally, researchers have sequenced convenience samples collected as part of routine clinical or public health activities (e.g., diagnostic specimens collected as part of an outbreak response), or as part of studies where specimens are collected for other purposes. However, the analysis of pathogen genomic sequences is increasingly becoming a primary goal of both research studies and public health surveillance efforts (Gardy et al. 2011; Jackson et al. 2016; Quick et al. 2016; Snider et al. 2016). This shift has been driven by the apparent utility of pathogen sequence data for understanding aspects of pathogen spread ranging from the frequency and source of introductions into a region (Nelson et al. 2007; Lei and Shi 2011; Thézé et al. 2018; Weill et al. 2019; Gonzalez-Reiche et al. 2020), to identifying endogenous spread of emerging diseases (Carroll et al. 2015; Park et al. 2015), to understanding the role of “hotspots” in maintaining broader community epidemics (Ratmann et al. 2020), to understanding transmission patterns at an individual or “microscale” level (Gardy et al. 2011; Salje et al. 2012).

Despite these many examples, there is a lack of clear and accessible guidance for appropriately designing and sizing studies aimed at understanding pathogen transmission, or for evaluating the design and conclusions of past studies. Without such guidance, it is difficult for researchers to design studies in a way that maximizes the chances of success, and difficult for reviewers to appropriately evaluate papers and grant applications centered around molecular or phylogenetic outcomes (Volz and Frost 2013; Frost et al. 2015). In particular, undersampling or biased sampling can lead to poorly supported inferences about patterns of disease spread (Grabowski and Lessler 2017; Mavian et al. 2020). While there are examples of researchers conducting careful *a priori* analyses of sampling strategies (Network and Others 2013; Farhat et al. 2014; Kelly et al. 2015), these have largely relied on sophisticated techniques that are not broadly generalizable. Hence, there is a need for broadly accepted and accessible guidance for the selection of specimens for sequencing and phylogenetic analyses.

As noted above, pathogen sequences have been used to understand multiple aspects of infectious disease transmission at scales ranging from the global (e.g., movement of pathogens between countries) to the individual (e.g., reconstruction of individual transmission chains). Arguably, all such analyses can be reduced to the basic question of whether pairs of infected individuals are related within a particular number of generations of transmission. Therefore, developing tools for assessing the number of sequences needed to confidently identify linked pairs (infections separated by no more than a specific number of generations of transmission) is a good place to start building a theory for power calculations for phylogenetic inference. In this paper, we present a framework for making critical decisions about study design when the goal is to identify infector-infectee pairs, and we illustrate this approach with simulation studies.

## Approach

### General Principles

In this paper we will deal with studies that aim to identify infector-infectee pairs from phylogenetic analysis of pathogen sequence data collected from infected individuals. We assume the study aims to achieve some level of certainty that identified infector-infectee pairs are correct, and may also require identification of some minimum number of pairs. Below we lay out a precise terminology (**Table 1**) and general principles.

**Table 1:** Parameters used in calculations and simulations.

| Parameter | Description |
| --- | --- |
| $M$ | Number of infections sampled |
| $N$ | Total number of (relevant) infected individuals in an outbreak |
| $\rho$ | Proportion of outbreak infections sampled ($M/N$) |
| $\eta$ | Sensitivity of the linkage criteria |
| $\chi$ | Specificity of the linkage criteria |
| $\phi$ | Probability that an identified link represents a true transmission event (1-False Discovery Rate) |
| $R$ | Reproductive number of a pathogen |
| $R_{\text{pop}}$ | Average reproductive number of a pathogen in a finite population (always $<1$) |
| $\mu$ | Mutation rate of the pathogen (in mutations per genome per transmission event) |

To start, we define the term *linkage criteria* to represent all the criteria used to infer whether a set of infected individuals are linked to one another by direct transmission. The *linkage criteria* can be derived from a combination of genetic distance between pathogens isolated from different individuals, tree structure (e.g., clade support), and epidemiologic information (e.g., relative dates of symptom onset). We refer to infections inferred to be connected by transmission using this criteria as *linked pairs*. Some of these linked pairs will represent actual transmission events (*true transmission pairs*) and some will be false positives. We want to determine the sample size (*M*) and proportion of the population (ρ) required to recover a predetermined number of linked pairs, while keeping the *false discovery rate* (the proportion of these linked pairs that are false positives) below a predetermined threshold. When applied to a study where design was dictated by other factors (e.g., specimen availability), the same methods can be used to determine the *false discovery rate*, which will inform the confidence we have in any conclusions about disease transmission in that study.

To capture *true transmission pairs*, the infector and their partner infectee must both be in the sample. Therefore, correctly identifying direct transmission links (and, conversely, calculating the false discovery rate) depends on the sampling fraction (ρ), which is equal to the sample size (*M*) divided by the total number of infected individuals in the relevant population (*N*). Identification of these links will further depend on the *sensitivity* (*η*) and *specificity* (χ) of the criteria used to define linkage. We define sensitivity as the probability that the linkage criteria will identify a true transmission pair as a linked pair given that both the infector and infectee are in the sample. Similarly, the specificity is the probability that two infections not linked by transmission are not linked by the linkage criteria. Here we show that, if we have reasonable estimates of the sampling fraction, sensitivity, and specificity, we can, for a sample of size *M*, estimate the false discovery rate. The relationship between these parameters can then be used to design studies with a sample size and sampling fraction that minimizes the *false discovery rate* and therefore maximizes our ability to draw inferences from identified infections.

### Calculating sample size and false discovery rate

#### Single link and single true transmission

We start with the simple example of identifying the correct infector of a particular infection (Volz and Frost 2013). In this scenario, we make assumptions about transmission that simplify the relationship between sample size and false discovery rate. Namely, we assume that each infected individual is connected by transmission to exactly one other individual, and that the linkage criteria similarly identifies exactly one probable link for each infection. Under these assumptions, we can calculate the probability of correctly identifying a true transmission pair, *ϕ*(equal to one minus the false discovery rate), as a function of the sensitivity and specificity of the linkage criteria, the proportion sampled, and the sample size. **Figure 1** provides some intuition as to the form of this probability expression under the stated assumptions of single linkage and single transmission (see **Text S1** for full derivation).

**Figure 1.**
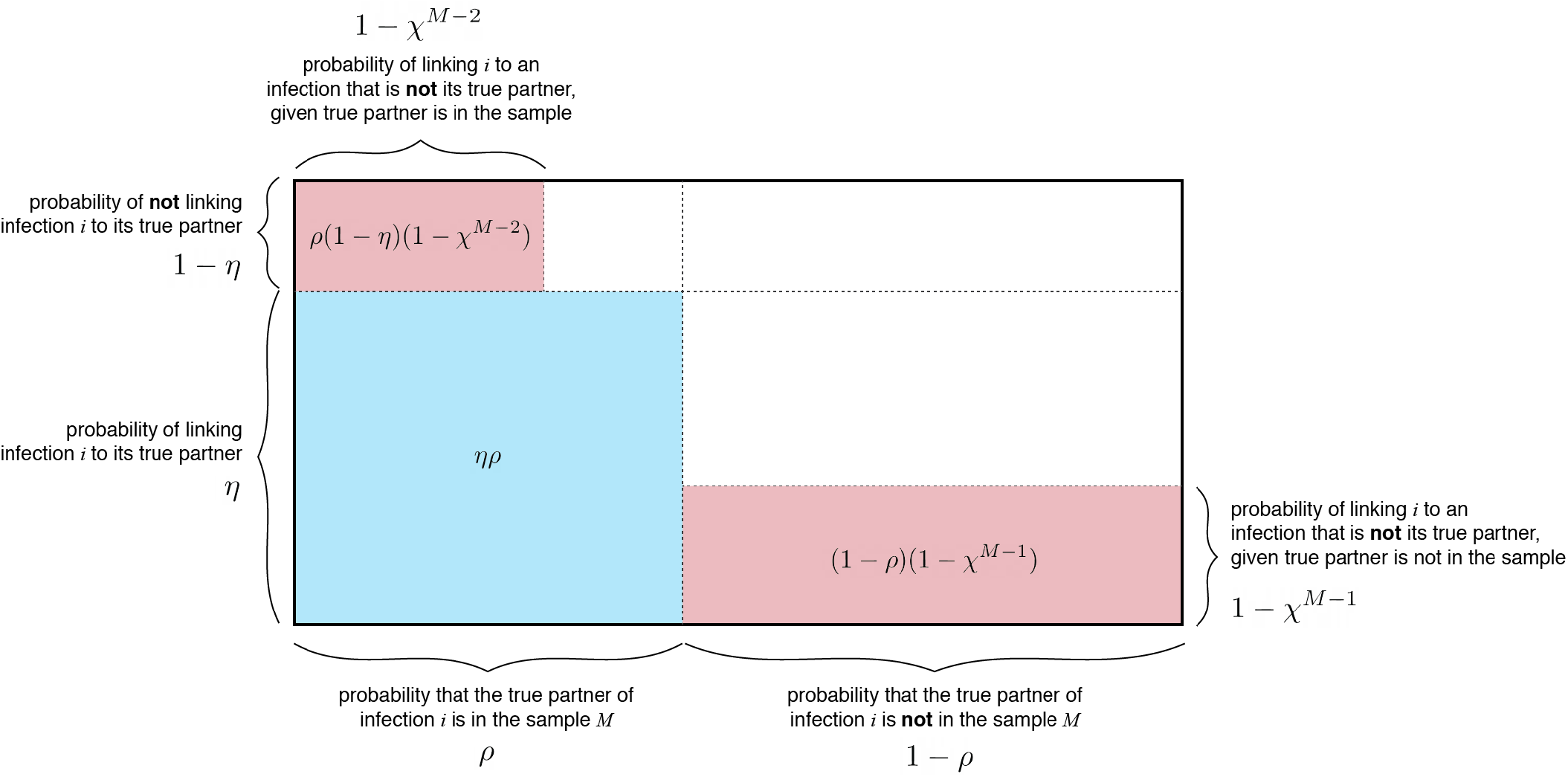
Visual derivation of the probability of correctly identifying a true transmission pair. Blue shaded regions represent correct identification of the true transmission partner of a random infection *i*. Red shaded regions represent linkage of *i* to an infection that is not its true transmission partner. White shaded regions represent the probability of no linkage occurring.

The probability of correctly identifying a true transmission pair (*ϕ*) under the assumptions of single transmission and single linkage is:

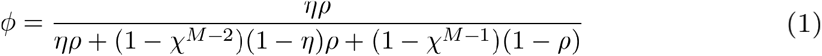

Under the same assumptions, we can also calculate the expected total number of true transmission pairs that will be identified in our sample, 𝔼[number of true pairs], as:

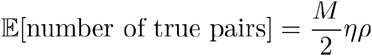

Through algebraic rearrangement of these equations, we can determine the expected number of linked pairs (identified with the linkage criteria) observed in this sample (𝔼[number of pairs observed]):

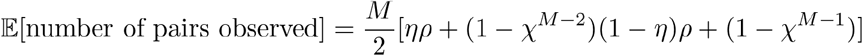

These equations can be used to determine the false discovery rate (1-*ϕ*) and the expected number of linked pairs given a particular criteria, sample size, and sampling proportion. Additionally, we can use these equations to observe how the expected number of links and the true discovery rate vary with the proportion sampled and the sample size (**Fig 2A**). For a given sensitivity and specificity of the linkage criteria, we observe that the false discovery rate *increases* with sample size if the proportion sampled remains constant, suggesting that studies aimed at correctly identifying the highest proportion of transmission links should prioritize sampling proportion over number of samples. Additionally, the relationship between false discovery rate and sampling proportion is dependent on the sample size needed to obtain that sampling proportion such that the impact of sampling proportion increases with sample size. We also observe the effects of changing sensitivity and specificity on the false discovery rate and find that the specificity of the linkage criteria is of key importance when attempting to minimize the false discovery rate of transmission pairs (**Fig 2B**).

**Figure 2.**
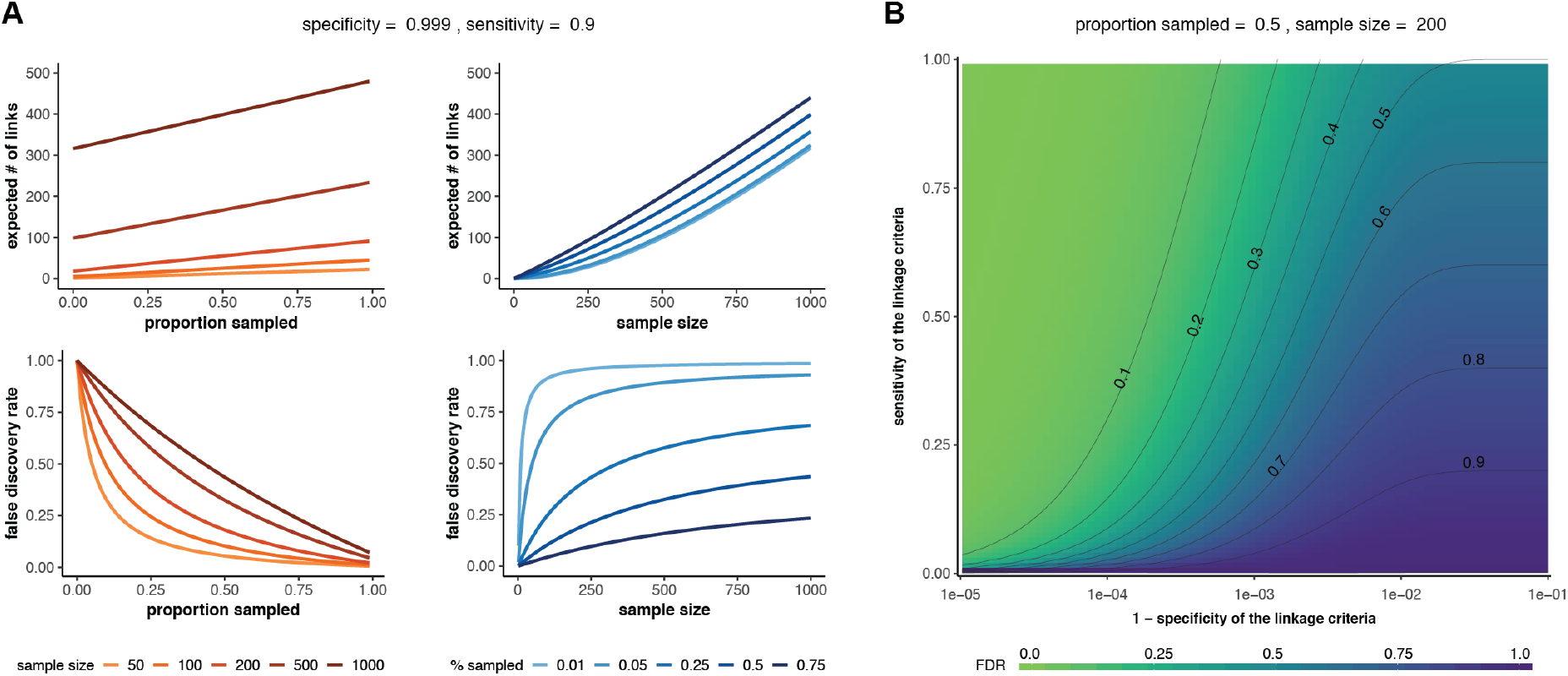
Sample size and false discovery rate given single linkage and single transmission. (**A**) Effect of sample size (red lines) or proportion sampled (blue lines) on the expected number of linked pairs (upper plots) or the false discovery rate of linked pairs (lower plots). The specificity and sensitivity are held constant. (**B**) Effect of varying the sensitivity and specificity of the linkage criteria on the false discovery rate (FDR).

#### Multiple links and multiple true transmissions

In many cases, we will be interested in linking an infected individual to both their infector and anyone they infect. Therefore, we must account for the fact that each infection in an outbreak may be linked by transmission to multiple other infections, only some of which may have been sampled. If the goal is to identify all such true transmission pairs in the sample, the linkage criteria used must similarly allow for multiple linkages. Here, we calculate the false discovery rate for transmission pairs under these assumptions.

The average number of transmission links per infection is determined by the epidemiological parameter *R*, the expected number of other individuals each infected individual infects. However, sampled infections come from a bounded source population. In this finite sampling frame, the average number of infectees per infector, denoted *R*_pop_, may differ from *R* (in fact, *R*_pop_ must be less than 1, see below). Because each infection is linked to, on average, *R*_pop_ infectees as well as its infector, each infection has *R*_*pop*_+*1* true transmission partners. If we assume that the distribution of the number of transmission partners per infection is Poisson distributed, we get the following equation for the true discovery rate, *ϕ*(see **Text S1** for full derivation):

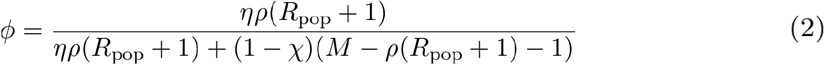

Under the same assumptions, we can calculate the total number of sampled true pairs, 𝔼[number of true pairs], as:

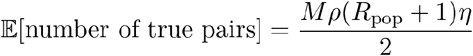

Through algebraic rearrangement of these equations we can determine the expected number of pairs observed in this sample, 𝔼[number of pairs observed]:

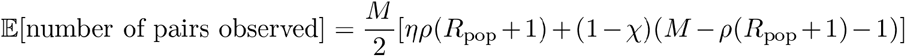

Again, we observe that the false discovery rate increases with the sample size, but decreases with the proportion sampled, and we see the important effect of the specificity of the linkage criteria on the false discovery rate (**Fig 3**).

**Figure 3.**
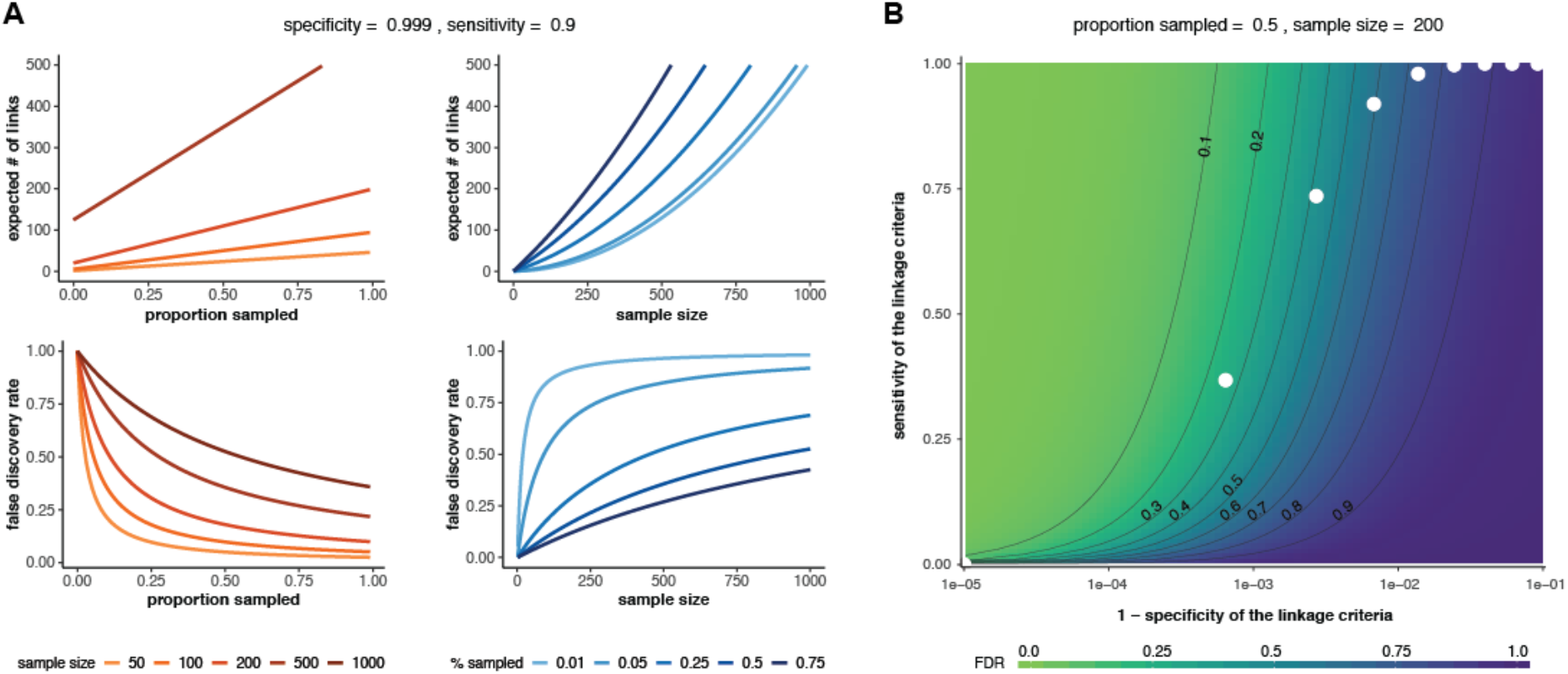
Sample size and false discovery rate given multiple linkage and multiple transmissions. (**A**) Effect of sample size (red lines) or proportion sampled (blue lines) on the expected number of linked pairs (upper plots) or the false discovery rate of linked pairs (lower plots). The specificity and sensitivity are held constant. (**B**) Effect of varying the sensitivity and specificity of the linkage criteria on the false discovery rate (FDR). White dots: theoretical sensitivity and specificity values at different genetic distance thresholds for a hypothetical pathogen with mutation rate = 1 mutation/genome/transmission and *R*=2 (see ‘Determining sensitivity and specificity’ below for details). In both panels, 𝔼_*pop*_=1.

### Estimating the average reproductive number

In the previous section, we distinguished *R*, the basic reproductive number of a pathogen, from *R*_pop_, the *average* reproductive number in a bounded population. This is an important distinction because we can show that the average reproductive number (*R*_pop_) is at most one. This is because any sampling frame contains a finite number of infected individuals. Therefore, there will always be more infections than infection events (at minimum, all infectees in a transmission chain plus a single index case, see **Fig S1**). Hence, *R*_pop_, which is equal to the number of actual transmission events divided by the number of infections, will be at most one.

In epidemic situations where there is a single introduction, *R*_pop_ will be close to one, as the number of infections will exceed the number of infection events by precisely one. In situations where there are multiple introductions (e.g., transmission chains that are persistently seeded from sources outside the sampling frame) then *R*_pop_ may be substantially less than one. Specifically:

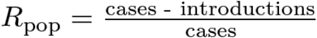

The examples shown in this paper focus on epidemics seeded by a single introduction, where *R*_pop_ is approximately equal to one.

### Determining sensitivity and specificity

In the framework presented here, the sensitivity and specificity of the linkage criteria are needed to estimate the false discovery rate from sample size and vice versa. This criteria can be based on a number of phylogenetic and epidemiological metrics, and may depend on the data available for a particular study. In this section, we outline two methods for approximating the sensitivity and specificity of a simple genomic metric: genetic distance.

Both methods involve determining these parameters from the discrete distributions of genetic distances between linked and unlinked infections, but they differ in how these distributions are obtained. Given the distributions, we can consider a number of different genetic distance thresholds (e.g., 2 mutations between sequences) that could be used as the criteria for differentiating between linked and unliked pairs, and we can calculate the sensitivity and specificity at each. The optimal threshold and its associated sensitivity and specificity can be selected in a variety of ways (Youden 1950; Perkins and Schisterman 2006; Liu 2012; Zou et al. 2013) based on the specific study goals.

Below, we describe two ways to obtain the genetic distance distributions of linked and unlinked infection pairs for a hypothetical pathogen with *R=2* and a mutation rate (*μ*) of 1 mutation per genome per generation. Here and henceforth, “generation” refers to a generation of transmission (i.e., the mutation rate provides the number of mutations expected per transmission event, not per viral replication).

#### Empirical method

One way to estimate the relevant genetic distance distributions is to use existing data. Specifically, we need a subsample of infections for which sequencing data is available and we have a high degree of confidence—based on epidemiological data—of the true transmission relationships between included infections. For example, infected individuals who share a household versus community members with no known relationship. We can compute the genetic distance between every pair of pathogen sequences from this subsample and use the results to approximate the underlying genetic distance distributions between linked and unlinked infections in the population.

We illustrate this method on a simulated outbreak of approximately 1500 infections (data available at https://github.com/HopkinsIDD/phylosamplesize), created using the *outbreaker* R package (R Core Team 2013; Jombart et al. 2014) (see ‘Outbreak simulations’ below). To create our known subsample, we selected a small number of infections from early in the outbreak and extracted their true transmission links and simulated genomes. We then calculated the genetic distance matrix of sequences in this subsample and determined the genetic distance distributions (**Fig 4A**). Next, we estimated the sensitivity and specificity at every mutation threshold (0 mutations, 1 mutation, etc.) and used the point closest to the (0,1) corner to determine the optimal threshold for differentiating between linked and unliked infections. In this case, the optimal threshold was 3 mutations, which had a sensitivity of 0.95 and a specificity of 0.88.

**Figure 4.**
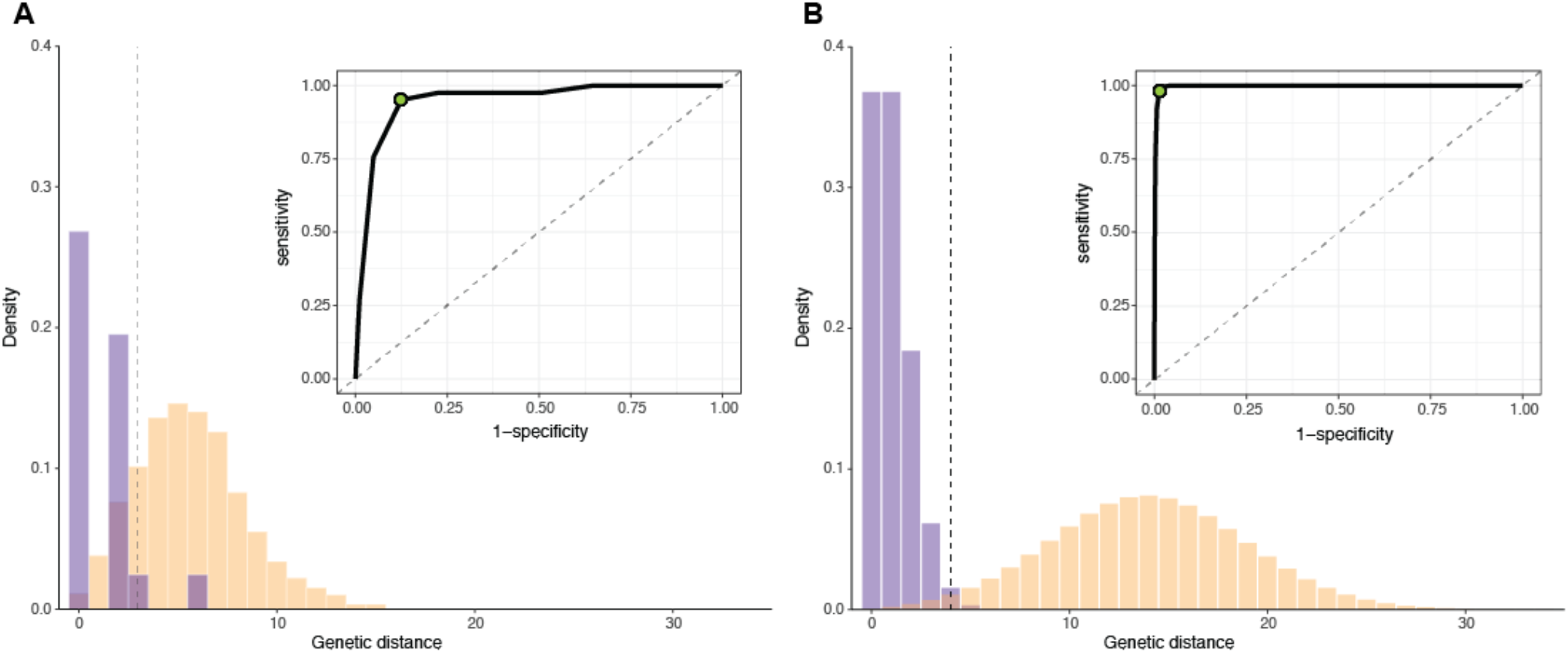
Determining the sensitivity and specificity of a genetic distance threshold. (**A**) Empirical distribution of genetic distances for linked (purple) and unlinked (yellow) infections for 50 infections selected from early in a simulated outbreak (*ϕ*= 1 mutation/genome/generation, R=2). Inset: receiver operating characteristic (ROC) for all possible genetic distance thresholds. Optimal threshold shown as green dot (ROC) and dashed vertical line (distribution). (**B**) Estimated distribution of genetic distances for linked and unlinked infections generated by the mutation rate method. Parameters and plots are as in (A).

#### Mutation rate method

Pathogen mutation rates can also be used to estimate the genetic distance distributions, especially when a subsample of infections with known transmission histories is not available. If we assume that the number of mutations between two linked infections is Poisson distributed around the mutation rate and that we know the distribution of the number of generations between infections in the population, the probability of observing a specific genetic distance (*d*) between the sequences from any two infected individuals linked by transmission is:

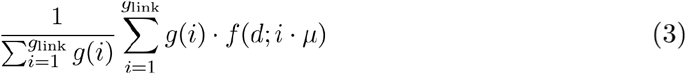

where *g*(*i*)is the probability of observing *i* generations between infections, *g*_*link*_ is the maximum number of generations between infections considered linked, *f*(*d*;*i*·*μ*)is the probability of observing *d* mutations between two infections separated by *i* generations, and *μ* is the mutation rate per genome per generation (see **Text S2**).

Similarly, the probability of observing a genetic distance *d* between two infections not linked by transmission is:

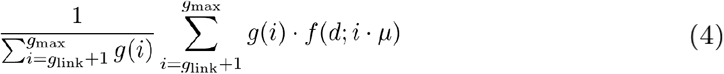

Where *g*_max_ is the maximum number of generations considered.

Determining the distribution of generations between infections is a non-trivial task (Dobrow 1996; Mahmoud and Neininger 2003; Salje et al. 2016), and depends on several factors, including the shape of the epidemic and the period of time from which infections are sampled (**Fig S2**). In the examples included herein, we use simulations to empirically approximate this distribution (see **Text S2**), but it is likely that adequate approximations can be obtained by other means—or that more sophisticated approaches can be employed to directly estimate the necessary genetic distance distributions (Worby et al. 2014).

Given the approximate generation distribution between infections, we calculated the genetic distance distributions for linked and unlinked infections for the pathogen described above. The optimal genetic distance threshold for distinguishing between linked and unlinked infections was 4 mutations (sensitivity=0.98, specificity=0.99) (**Fig 4B**). The empirical and mutation rate methods result in a similar, but not identical, optimal threshold for the pathogen in this example, likely due to sparse sampling in the empirical case.

Additionally, we note that the clear threshold (and high sensitivity and specificity) observed here only occurs when the mutation rate is high enough (and the reproductive number low enough) that a significant number of mutations occur between infections considered linked (Campbell et al. 2018). For pathogens that do not meet these criteria, it may not be possible to use genetic distance alone to distinguish between linked and unlinked infections (**Fig S3**).

## Methods

### Outbreak simulations

We used outbreak simulations to validate our approach. We simulated outbreaks using the ‘simOutbreak’ function implemented in the *outbreaker* R package (Jombart et al. 2014). For all simulations we assumed a large number of susceptible individuals in the population (n.hosts=100,000), a genome length of 1,000 nucleotides, and no importation events (single source outbreak). We also assumed every infected individual transmitted their infection exactly one time step after infection, and ran the simulation for the number of generations needed to achieve a final outbreak size of approximately 1,000 infections (*In*(*1,000*)/*In*(*R*)). After simulating the source population, we randomly selected a predetermined proportion of infections from that population.

For each sampling proportion, we simulated outbreaks over a variety of mutation rates and reproductive numbers. We allowed the mutation rate to vary between 0.0001–4 mutations per genome per generation, and allowed the reproductive number to vary between 1.3–18. We chose these ranges to encompass mutation rates and reproductive numbers observed in actual human pathogens. We divided each parameter range into 100 discrete values and ran simulations with all combinations of mutation rate and reproductive number, for a total of 10,000 simulations for each sampling proportion. We required simulated outbreaks to contain at least 100 and no more than 2000 infections.

### Implementation

Functions for calculating the necessary sample size based on a desired false discovery rate are implemented in the R package *phylosamp*, freely available at: https://github.com/HopkinsIDD/phylosamp. This package also includes functions for calculating the false discovery rate for a specific sample size or proportion, and functions to estimate the number of transmission pairs that will be observed given a sample size and a set of assumptions (e.g., multiple links and multiple transmissions, single link and single transmission, etc.). We also provide generation distributions for values of *R* between 1.3–18, derived from the simulations described in **Text S2**.

## Results

### Method performance with known sensitivity and specificity

We used simulated outbreaks to validate the relationship between sample size and false discovery rate using genetic distance as our linkage criteria. We subsampled each outbreak and, using the known transmission relationships and genetic distances between simulated infections, calculated the false discovery rate at each possible genetic distance threshold in the subsample (“simulated FDR”). For each simulation (before subsampling), we also calculated the actual specificity and sensitivity at every relevant genetic distance threshold. We used these values and the observed *R*_pop_ (roughly equal to one in most simulations) to then calculate the theoretical false discovery rate at a particular sampling proportion using **Equation 2**. We find that the theoretical false discovery rate is consistent with the simulated value for a wide array of pathogen mutation rates and reproductive numbers (**Fig 5**).

**Figure 5:**
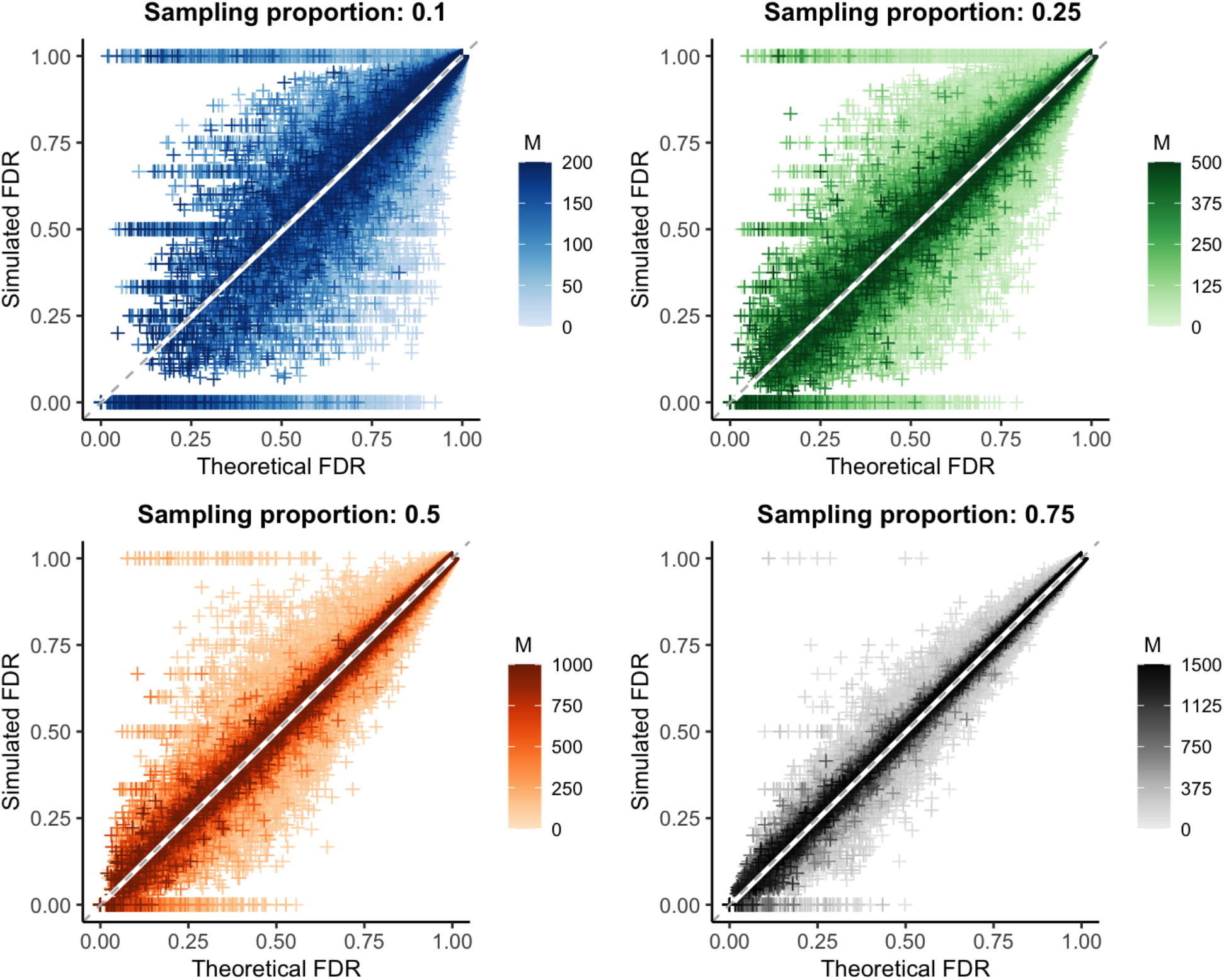
Predicted versus observed false discovery rate in outbreak simulations. Theoretical versus simulated false discovery rate (FDR) for each genetic distance threshold in 10,000 simulations of varying mutation rate and reproductive number. White line: smoothed conditional mean; grey dashed line: y=x line. Increasing values of the sample size (*M*) are plotted in darker color; because the maximum outbreak size is fixed at 2000, the maximum sample size differs for each sampling proportion. Increasing both the sample size and proportion reduces bias and error, see **Table 2**.

Overall, the bias of our estimate of the false discovery rate approached zero for all sampling proportions. The average error was less than 4% in each case, decreasing significantly with increased sample size or proportion sampled (**Table 2, Table S1**). We note that special care should be taken with low sample sizes and low theoretical false discovery rates, as error rates can be particularly high in this range. Additionally, while our method is an unbiased estimator and overall correct in expectation, it is always possible for performance in a particular set of individuals sampled from a population to deviate substantially from expectation (for example, when a subsample happens to contain no true transmission pairs), particularly when sample sizes are low.

**Table 2:** Bias and error of calculated false discovery rate for simulations with fixed sampling proportion.

| Bias | $\rho=0.10$ | $\rho=0.25$ | $\rho=0.50$ | $\rho=0.75$ | All $\rho$ values | N |
| --- | --- | --- | --- | --- | --- | --- |
| FDR=0.00-0.25 | -0.0006 | 0.0045 | 0.0001 | 0.0036 | **0.0022** | 17,900 |
| FDR=0.25-0.50 | 0.0044 | 0.0045 | 0.0009 | 0.0032 | **0.0032** | 31,633 |
| FDR=0.50-0.75 | 0.0064 | 0.0039 | 0.0006 | 0.001 | **0.0029** | 51,069 |
| FDR=0.75-1.00 | 0.0001 | 0.0001 | <0.0001 | <0.0001 | **0.0001** | 965,125 |
| All FDR Values | **0.0005** | **0.0005** | **0.0001** | **0.0002** | **0.0003** | 1,065,727 |
| N | 261,360 | 267,239 | 268,900 | 268,228 | 1,065,727 |  |

| Error | $\rho=0.10$ | $\rho=0.25$ | $\rho=0.50$ | $\rho=0.75$ | All $\rho$ values | N |
| --- | --- | --- | --- | --- | --- | --- |
| FDR=0.00-0.25 | 0.2135 | 0.1359 | 0.0799 | 0.0401 | **0.098** | 17,900 |
| FDR=0.25-0.50 | 0.2751 | 0.1583 | 0.079 | 0.0416 | **0.1275** | 31,633 |
| FDR=0.50-0.75 | 0.2057 | 0.0979 | 0.0478 | 0.0259 | **0.092** | 51,069 |
| FDR=0.75-1.00 | 0.0155 | 0.0069 | 0.0035 | 0.002 | **0.007** | 965,125 |
| All FDR Values | **0.032** | **0.0181** | **0.0097** | **0.0052** | **0.0161** | 1,065,727 |
| N | 261,360 | 267,239 | 268,900 | 268,228 | 1,065,727 |  |

To better understand why the error rate of our estimator increases as the false discovery rate decreases, we stratified the simulation data by the sensitivity and specificity given a particular genetic distance threshold. We found that the error is highest when sensitivity is low and specificity is high (**Fig S4A-B**), which occurs when a high genetic distance threshold is used. This combination often produces low false discovery rates, but is highly dependent on sampling (namely, if any true positives or false positives are sampled). This leads to highly variable simulated false discovery rates and consequently higher error rates. Unsurprisingly, this analysis also highlights that a discrete threshold like genetic distance produces a limited number of possible sensitivity and specificity combinations (**Fig S4C-D**). Therefore, obtaining reasonable estimates for these values in tandem is of key importance when using our method to estimate the false discovery rate of a phylogenetic study.

### Method performance with estimated sensitivity and specificity

We repeated the false discovery rate comparison described above, but instead of using the actual sensitivity and specificity observed in each simulation, we calculated these parameters from the mutation rate used to generate that simulated outbreak (**Fig 6**). To reduce reliance on simulation data to calculate necessary parameters, we used *R*_*pop*_=*1*, rather than the empirical value.

**Figure 6:**
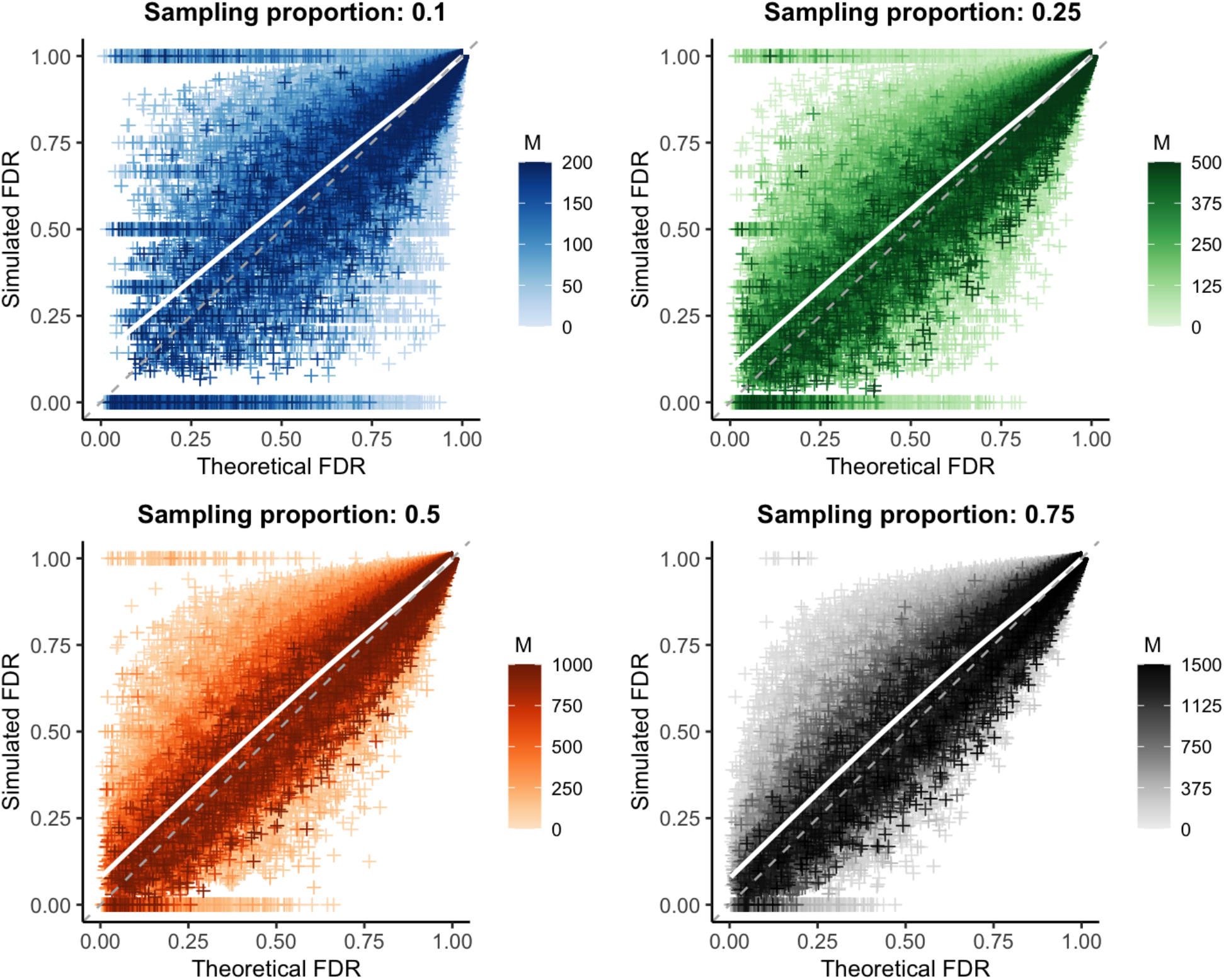
Validation of mutation rate method to calculate sensitivity and specificity. Theoretical versus simulated false discovery rate (FDR) for each genetic distance threshold in 10,000 simulations of varying mutation rate and reproductive number. White line: smoothed conditional mean; grey dashed line: y=x line. Increasing values of the sample size (*M*) are plotted in darker color; increasing both the sample size and proportion reduces bias and error, see **Tables S2** and **S3**.

Under this more realistic set of assumptions, we observe a slight bias, though overall values remain less than one percent (**Table S2, Table S3**). However, while mean bias is very low on average, it is greater when the theoretical false discovery rate is low, reaching nearly 8% for predicted false discovery rates less than 25%. Average error rates were similarly slightly increased, but remained less than 4% overall.

Given that correct sensitivity and specificity values are an important component of calculating the theoretical false discovery rate, we looked at the specific estimates for these parameters generated by our mutation rate method. When considering only direct transmissions as linked (as we do throughout these simulations), **Equation 3** simplifies to simply a poisson distribution around the mutation rate, resulting in highly accurate and precise sensitivity estimates (**Fig S5**). However, we find that our estimates for specificity (**Fig S6**) have some positive bias (and large error, particularly for low sample sizes). We hypothesized that inaccuracies in the estimated specificity were due to the distribution of generations between infections used in our calculation; as discussed in **Approach**, this is a non-trivial distribution that we estimated by averaging over many simulations (see **Text S2** for details).

To test this hypothesis, we used the actual distribution of generations between infections from each simulation in our calculation of specificity (sensitivity estimates are unaffected by this distribution when considering only direct transmissions, as described above). We find that this does in fact reduce bias in our specificity estimates (**Fig 7**) and leads to largely unbiased (<2%) estimates of the false discovery rate, even at low theoretical false discovery rate values (**Fig S7, Table S4**).

**Figure 7:**
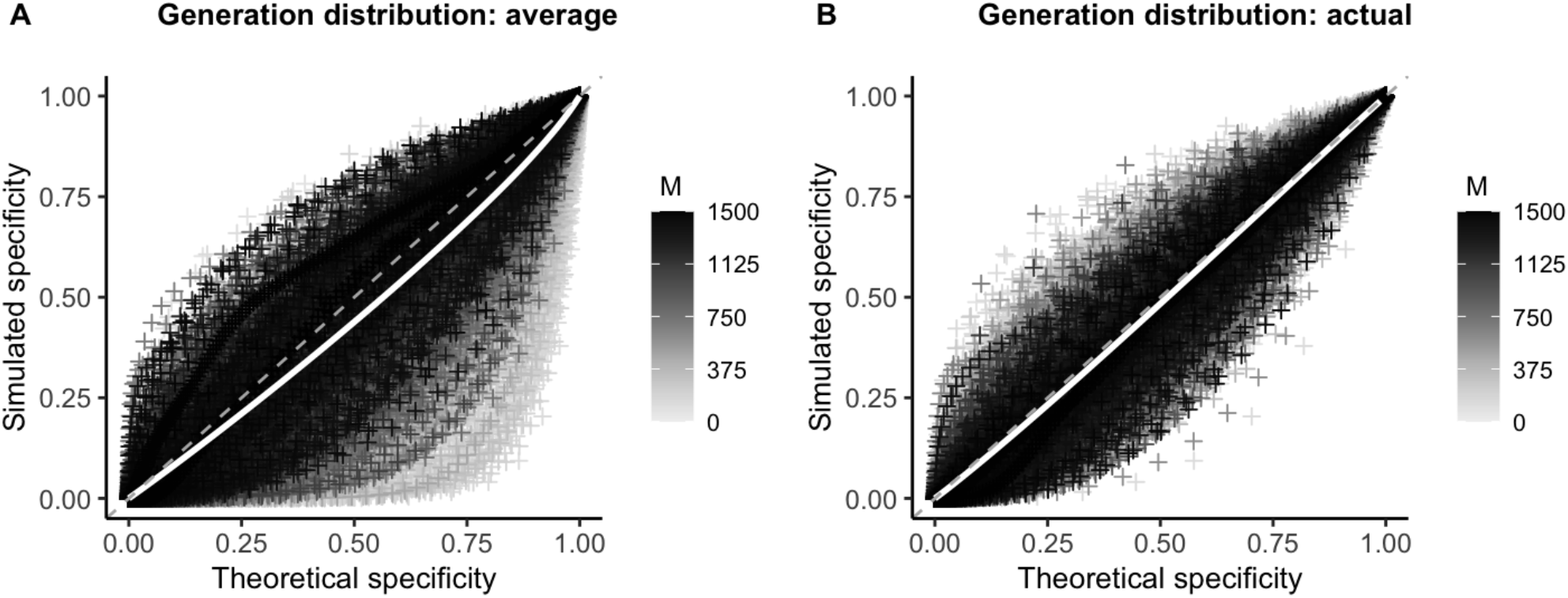
Effect of the generation distribution on specificity of the linkage criteria. Theoretical versus simulated specificity for each genetic distance threshold in 10,000 simulations of varying mutation rate and reproductive number (proportion sampled = 0.75). White line: smoothed conditional mean; grey dashed line: y=x line. Increasing values of the sample size (*M*) are plotted in darker color. (**A**) Theoretical sensitivity and specificity calculated using average distribution of generations between infections from simulations (see **Text S2**). (**B**) Theoretical sensitivity and specificity calculated using the actual distribution of generations between infections from that simulated outbreak.

## Discussion

We have developed a mathematical framework for making informed sampling decisions in pathogen genome sequencing studies. Specifically, this framework allows for easy calculation of the relationship between the number or proportion of infections sampled during an outbreak and the ability of some phylogenetic or epidemiological criteria to correctly identify infections within this sample that are linked by direct transmission. Understanding this relationship is crucial to making correct inferences about pathogen transmission patterns, especially as genomic studies are becoming more feasible and widely used to answer both scientific and public health questions.

This framework is broadly applicable to a variety of phylogenetic or epidemiological approaches, as long as the sensitivity and specificity of the criteria can be approximated. With a basic understanding of the pathogen and the criteria being used, researchers can more effectively design studies that correctly identify transmission pairs with a known level of confidence. Additionally, this generalizable method (available as a free software, the R package *phylosamp*) provides a metric by which reviewers of these studies can evaluate their conclusions. We apply our method to simulated outbreaks using genetic distance as the linkage criteria and find that we can effectively estimate the false discovery rate for a variety of pathogen mutation rates, reproductive numbers, and relevant genetic distance thresholds. It is important to note, however, that for a given sensitivity and specificity, there may not always be a study design that achieves the desired false discovery rate.

Performance of the method presented depends on our ability to estimate the sensitivity and specificity of a particular linkage criteria. While we present two methods for doing this—empirically and theoretically using the mutation rate of the pathogen—implementing either in practice is not without challenges, and improved estimation of these values may be a fruitful area for future research. For instance, the mutation rate based approach also depends on the distribution of the number of generations of transmission between infections in the underlying population. Although distributions derived from simulations (provided as part of the *phylosamp* package) provide a reasonable proxy, estimates of sensitivity and specificity are much improved when using the exact generation distribution, which currently can only be determined from complete knowledge of all transmission events. Further research into all the factors affecting this distribution will be necessary to improve its estimation. Likewise, there are challenges to the empirical approach, particularly for novel pathogens.

Better performance can likely be obtained by not restricting ourselves to genetic distance alone when determining a linkage criteria. Genetic distance is easy to determine from sequence data, but this simple metric does not take into account ancestral relationships or uncertainty around these relationships, and is limited to discrete mutational changes. Applying more complex phylogenetic criteria may allow us to learn more about transmission relationships, though there is a limit to the extent to which genetic data can be used to distinguish infections in fast-spreading (or slow-mutating) pathogen outbreaks. There are several examples of outbreaks in which multiple infected individuals have the same consensus viral genome (Campbell et al. 2018). In this case, incorporating epidemiological data (e.g., location, time of symptom onset) may be important in determining which infections are unlikely to be linked. Doing so is part of a larger effort to better integrate epidemiological and genomic data into pathogen transmission studies (Morelli et al. 2012; Ypma et al. 2012; Jombart et al. 2014; Klinkenberg et al. 2017).

While in this manuscript we have focused on direct transmission pairs, our framework is designed to be extensible to alternative definitions of linkage; for example, infections connected within a specified number of transmission events. Expanding the definition of linkage to include such indirect transmissions has a number of useful applications in outbreak research, such as identifying and connecting transmission clusters. This method could also be extended to more complex direct transmission relationships, for example when within-host evolution results in the existence of viral quasispecies within infected individuals, each of which has some potential of being transmitted. In all of these scenarios, it is equally important to understand the sample size needed to make the desired inferences.

We hope that this work represents a step towards developing a larger theory of study design for making inferences from pathogen sequence data, but recognize it is only a step. The focus of this paper is sample size, but which infections are sampled may be equally important (Stack et al. 2010; de Silva et al. 2012; Hall et al. 2016). For example, understanding routes of direct transmission may require dense sampling of a small group of highly-connected individuals, while understanding general transmission trends over the course of a geographically-ispersed outbreak may require us to sample broadly over space and time. Additionally, the goal of linking infections is seldom the linkages themselves, but the larger inferences about risk and transmission derived from those linkages. Adapting the techniques here to more directly link sample size calculations to these outcomes is an important next step.

## Data Availability

All simulations and code used as a part of this manuscript are publicly available on github.

https://github.com/HopkinsIDD/phylosamplesize

https://github.com/HopkinsIDD/phylosamp

## Acknowledgements

We thank Stuart Ray for his insightful comments on the manuscript. Funding was provided by Bill and Melinda Gates Foundation OPP1195157 (S.W. and J.L.).

